# Early life adversity and late life dementia in the Harmonised Cognitive Assessment Protocol network (U.S., China, England and Europe)

**DOI:** 10.1101/2025.06.04.25328976

**Authors:** Gindo Tampubolon, Guanan Li

## Abstract

Social progress and medical advances since the last century have raised uneven global challenges. The Lancet commission on dementia released a recent estimate and projection of dementia: more than 57 million older adults today and nearly triple that number in 25 years, mostly in developing nations. Its life course frame is expansive on modifiable risk factors in mid and late life but restrained on early life.

**MATERIALS AND METHODS:** We rectify this omission, assembling harmonised cognitive assessments from wealthy and developing nations (HCAP network) matched with retrospective childhood information obtained from 12,862 adults aged 69 years on average and 52% female. Error-laced recollection of early life adversity was analysed using latent construction and its association with cognitive status (normal, mild cognitive impairment/MCI and dementia) was estimated using multinomial logistic regression. Extensive sensitivity analyses were conducted. We plotted marginal probabilities of MCI and dementia for various groups.

**RESULTS AND DISCUSSION:** Harmonised prevalence rates of MCI and dementia vary considerably. Early life adversity is found to be associated with late life MCI and dementia in England and Europe. Not so in China and U.S. where selective mortality throughout the life course in largely private health care systems may have played a prominent role. In England and Europe the relative risks of early life adversity on MCI are 1.50 (95% confidence interval 1.16 – 1.92) and on dementia 1.52 (1.12 – 2.06).

These results uncover another manifestation of the life course shaping of health around the world, evidence made possible by harmonised methodologies which emerge as handy allies in global initiatives such as the UN Decade of Healthy Ageing. The analysis demonstrates that it is never too early to invest in healthy ageing.

## INTRODUCTION

The *Lancet* Commission on Dementia in 2024 estimated 57 million people with dementia worldwide just before the covid-19 pandemic and projected that in 25 years the number will nearly triple, mostly in developing nations.^1^ It has estimated the cost to be USD818 billion with 85% borne not by medical care but by families and societies. While age remains a major risk factor,^2–5^ the commission emphasised the hopeful message that modifiable risk factors across the life course are considerable, contributing to 45% of population attributable fraction.^1^ But compared to extensive mid and late life, the commission’s life course frame below has yet to unpack early life. We will begin to fill this omission.

**Figure 1.**
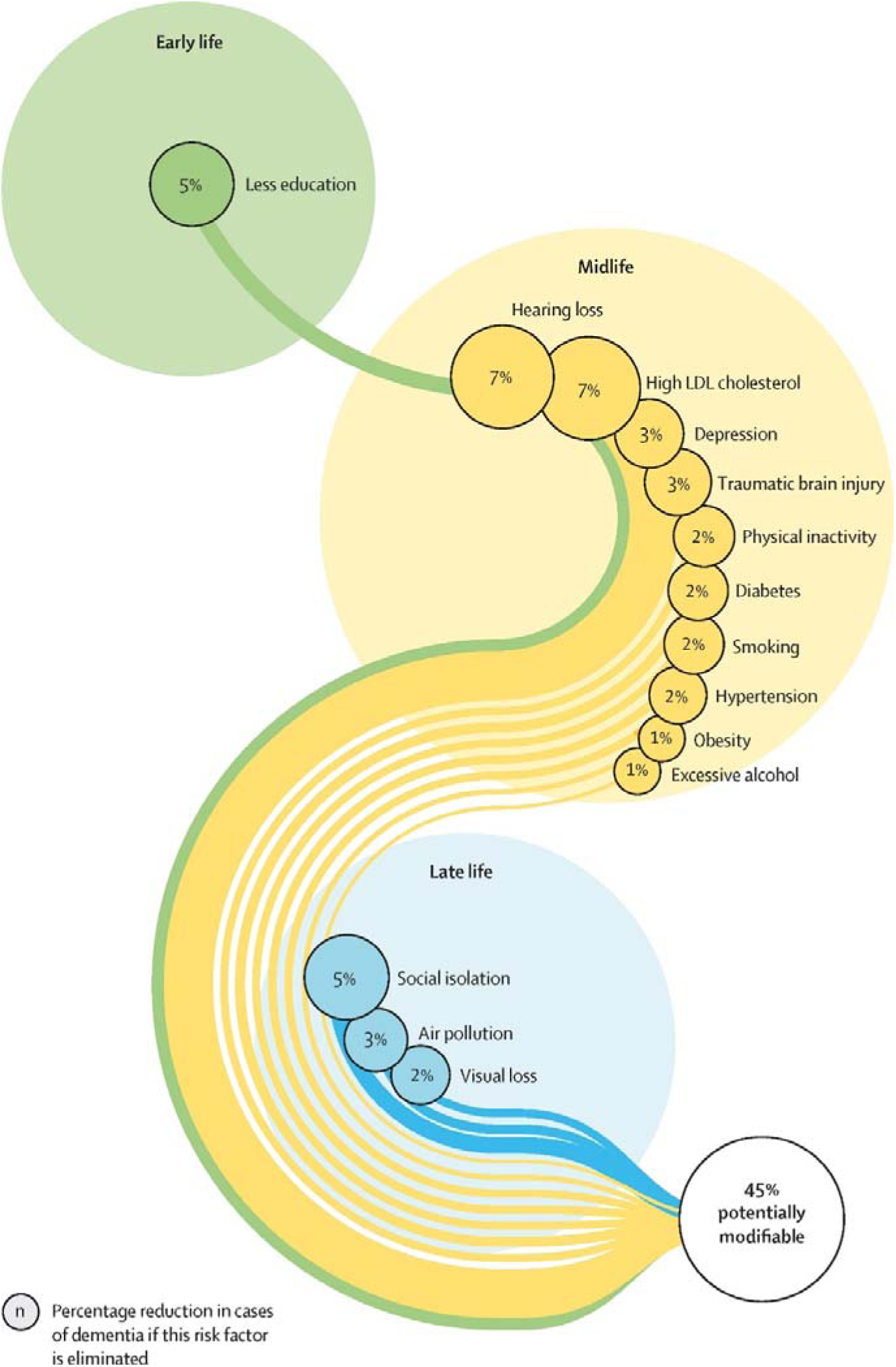
Lancet Commission on Dementia in late life – life course frame^1^

We aim to obtain new estimates of association between early life adversity and dementia in adults aged 50 and over while accounting for other covariates, a specific test of the life course shaping of health hypothesis.^6–18^ We do this for the largest number of countries (both wealthy and developing) using a harmonised cognitive assessment protocol, built on neurologically adjudicated instruments and algorithm, a protocol enjoying increasing adoption worldwide.^2,3,19–24^

Early life adversity is a significant risk factor for disability, dysfunction and disease in older adults in the United States (U.S.), Britain, China, Europe and Indonesia.^6–15^ This raises the question of whether early life adversity also predicts severe cognitive impairment or dementia, a condition associated with adverse clinical outcomes, raised medical costs, diminished life satisfaction and shortened life span.^1,4^ Evidence from 28 trans-Atlantic countries, including U.S., Britain and Israel, demonstrates persistent associations between early life adversity and key indicators of health in older adults, such as episodic memory, gait speed, sarcopenia, depression, frailty and multimorbidity.^7–10^ These are thought to operate, in part, through epigenetic mechanisms such as methylation.^8,25–28^ Understanding whether dementia in late life is also shaped by early life adversity is essential for informing global initiatives like the UN Decade of Healthy Ageing (accessed 16 March 2023).

Nations worldwide are experiencing population ageing (at least 7% of the population are 65 or over), with large developing nations including China and Indonesia ageing at a faster pace than historically experienced by wealthy nations. This presents a growing challenge: many countries are ageing before becoming wealthy. Because advanced age is a major risk factor for dementia,^1^ to ride the population transition we need to urgently understand its global prevalence and its other risk factors using harmonised methodologies.^3,19,20,23,24^ We ask: what are the harmonised prevalence rates of mild cognitive impairment (MCI) and severe cognitive impairment (dementia) in the U.S., China, England and Europe? After standard adjustment, does early life adversity remain associated with dementia?

To preview, our findings reveal substantial variation in the prevalence of MCI and dementia across these regions. However, despite differences in prevalence, the association between early life adversity and dementia persists. Adults who experienced early life adversity face higher risks of dementia later in life. Addressing early life adversity offers a lifelong benefit, potentially reducing the burden of cognitive decline across nations and generations.

## MATERIALS AND METHODS

### Data Sources

The Harmonised Cognitive Assessment Protocol (HCAP) is a set of standardised neuropsychological tests designed to assess cognitive function in ageing populations around the world. It is typically part of a larger longitudinal ageing survey. We used data from four of these: U.S. Health and Retirement Study (HRS), the China Health and Retirement Longitudinal Study (CHARLS), the English Longitudinal Study of Ageing (ELSA), and the Survey of Health, Ageing and Retirement in Europe (SHARE) – all part of the HRS-family. HCAP aims to provide comparable data on cognitive functions by harmonising cognitive assessments across diverse populations. Developed as part of the HRS-family, <u>HCAP</u> measures facets such as memory, executive function and language function (accessed 1 April 2025). Its standardised approach enables reliable comparisons of cognitive impairment across different populations. Stemming from the HRS, HCAP has been adapted to CHARLS, ELSA and SHARE i.e. Czechia, Denmark, France, Germany, Italy.^3,19,20,24^ The adaptation ensures cultural relevance and linguistic accuracy while maintaining the standard set, making cross-country comparisons meaningful for cognitive ageing. While HCAP is being adapted by many more countries such as in the Irish Longitudinal Study on Ageing, only HRS, CHARLS, ELSA and SHARE have earlier collected childhood adversity information, the key exposure. Participants with both early life information and HCAP information were retained. Note that although HCAP was harmonised, the collection of early life information was less so, leading to varying number of sample matchings between early life adversity and late life cognition across ageing surveys to no apparent bias in similar investigations.^8,9^

### Outcome: threefold cognitive status

The commission synthesised a wide range of evidence which used neuropsychological tests similar to HCAP.^4,29–31^ We closely followed HCAP methodology, algorithm and nomenclature as laid out by Jones and colleagues; also personal communication with Jones.^2,19,24^ Briefly, cognition is multifaceted: orientation, visuospatial, memory, executive function and language fluency. The participants’ test results are accompanied by informants’ reports of functions, namely Jorm and Blessed part I. Using the test results and the reports as input, following the HRS HCAP algorithm or flowchart,^2,19^ each participant is classified as normal or with mild cognitive impairment (MCI) or with severe cognitive impairment (dementia), nominal and mutually exclusive because not all MCI progresses to dementia.^32^ In this nomenclature, severe impairment status is equivalent to dementia or Alzheimer’s disease related dementia.^2,19,24^ In sum the threefold multinomial cognitive status is the outcome. For sensitivity analyses (1) the last two are collapsed, giving a binary outcome: normal or with any impairment, or (2) an ordinal cognitive status (normal, MCI, dementia) is used.

### Key exposure: Early life adversity or poverty

The participants were asked retrospectively about their childhood, for some this was eight decades earlier. (HRS: in 2015, 2017, 2019; ELSA: in 2006; CHARLS: 2014; SHARE: 2008-2009) Obtaining information on childhood adversity from adults with an average age of 69 year raises concern about recall errors. For example, in 2008 some 2500 Britons aged 50 were asked to recall the numbers of rooms and people in their homes when they were eleven to assess overcrowding (the British 1958 cohort).^33,34^ Only one in three got both numbers right. Their mothers gave the right answers when visited four decades earlier. Thus it is scarcely plausible that an average 69-year-old achieved an error-free recall of childhood. Assuming error-free early life information is unsafe.

Following prior practice of researchers from America, Britain and Europe which used latent constructs to address recall error, we constructed a binary latent class of childhood adversity (poor or non-poor).^6–10,16,18^ More important than unsafe inference, using recalled information unprocessed also prevents cross-country learning. For example, to explain the health of older Chinese both Si and colleagues and Li and colleagues used retrospective childhood conditions collected in CHARLS including death from starvation during China’s Great Leap Forward.^12,13^ But that information, unprocessed, complicates comparison with other countries’ childhood conditions because no government in the U.S. England, Czechia, Denmark, France, Germany or Italy ever imposed a comparable edict. Conversely, an indicator of the presence of running hot water in ELSA and SHARE will equally complicate the comparison because this was not asked in CHARLS or HRS. To overcome these, the researchers have devised a latent construct for early life adversity which enables it to be used as a risk factor to explain health in late life.^6–10,16–18^ The early life indicators in HRS, CHARLS, ELSA and SHARE which we used have been described elsewhere.^7–10^

### Covariates

Following the empirical literature on cognitive ageing this information is also included as covariates: marital status (unmarried/divorced/separated, married/union), residence (rural or urban, hukou is used in CHARLS), wealth quartiles and depression. Education is coded as a two-level variable: up to high school versus college or more (HRS, SHARE and ELSA); illiterate versus literate (CHARLS).

### Analysis

We fitted five multinomial logistic regressions: one for a pooled sample (HRS-HCAP, ELSA-HCAP, SHARE-HCAP, and CHARLS-HCAP), then one for each survey. Note that SHARE-HCAP warns against decomposing into five countries so we did not fit one for each country.^24^ We adjusted the standard errors for use of derived childhood adversity as the exposure following previous practice.^6–10,35^ To ease interpretation we produce plots of marginal probabilities across ages, distinguishing the childhood poor from the non-poor.

As per prior practice with HCAP all observations are used including those with incomplete items, imputing them using full information maximum likelihood or expectation maximisation algorithm.^2,8,19,24^ Latent class analysis was conducted with Latent Gold version 6^36^ and regressions with Stata version 19;^37^ significance level was set at 5 percent.

## RESULTS

We present a summary of the analytic sample from the eight countries in Table 1 which shows that 52% are female and its average age is 68.9 years. The largest cohort is from China followed by the US. Table 2 shows prevalence rates of normal, MCI and dementia. The prevalence across the countries suggests that the harmonisation was successful as shown by the stability of proportions across the developed countries on both sides of the Atlantic. Meanwhile, China’s result is apparently contradicting its middle-income context which might have led one to initially expect a higher prevalence of cognitive impairment. But this result is possibly driven by selective mortality or survival, commonly observed in population ageing.^14,24^ This suggests that, compared to wealthy countries, older people in China experienced earlier mortality for the same degree of impairment; conversely, in China only those with less impairment can survive to older ages, giving lower prevalence when a harmonised threshold was used. The regression results are collected in table 3.

**Table 1.** Summaries of HCAP matched with early life adversity information from four ageing surveys of HRS, CHARLS, ELSA & SHARE (N, %; mean, standard deviation)

|  | Male<br>N=6130 | Female<br>N=6732 | Total<br>N=12862 |
| --- | --- | --- | --- |
| Early life adversity |  |  |  |
| Non-poor | 5556 (90.6%) | 5948 (88.4%) | 11504 (89.4%) |
| Poor | 574 (9.4%) | 784 (11.6%) | 1358 (10.6%) |
| Cognitive status |  |  |  |
| Normal | 5415 (88.3%) | 5742 (85.3%) | 11157 (86.7%) |
| Mild or MCI | 568 (9.3%) | 776 (11.5%) | 1344 (10.4%) |
| Severe or dementia | 147 (2.4%) | 214 (3.2%) | 361 (2.8%) |
| Age | 68.9 (8.6) | 68.9 (12.0) | 68.9 (10.5) |
| Some college |  |  |  |
| No | 2688 (43.8%) | 4643 (69.0%) | 7331 (57.0%) |
| Yes | 3442 (56.2%) | 2089 (31.0%) | 5531 (43.0%) |
| Marital status |  |  |  |
| Unmarried/sep./divorced | 1202 (19.6%) | 2563 (38.1%) | 3765 (29.3%) |
| Married/union | 4928 (80.4%) | 4169 (61.9%) | 9097 (70.7%) |
| Wealth |  |  |  |
| Poorest quartile | 1384 (22.6%) | 1833 (27.2%) | 3217 (25.0%) |
| Higher | 4746 (77.4%) | 4899 (72.8%) | 9645 (75.0%) |
| Residence |  |  |  |
| Rural | 2271 (37.0%) | 2564 (38.1%) | 4835 (37.6%) |
| Urban | 3859 (63.0%) | 4168 (61.9%) | 8027 (62.4%) |
| Depression |  |  |  |
| No | 5338 (87.1%) | 5251 (78.0%) | 10589 (82.3%) |
| Depression | 792 (12.9%) | 1481 (22.0%) | 2273 (17.7%) |
| Country |  |  |  |
| China | 4352 (71.0%) | 4498 (66.8%) | 8850 (68.8%) |
| America | 539 (8.8%) | 735 (10.9%) | 1274 (9.9%) |
| England | 357 (5.8%) | 460 (6.8%) | 817 (6.4%) |
| Germany | 242 (3.9%) | 202 (3.0%) | 444 (3.5%) |
| Italy | 137 (2.2%) | 203 (3.0%) | 340 (2.6%) |
| France | 158 (2.6%) | 216 (3.2%) | 374 (2.9%) |
| Denmark | 214 (3.5%) | 207 (3.1%) | 421 (3.3%) |
| Czechia | 131 (2.1%) | 211 (3.1%) | 342 (2.7%) |

**Table 2.** Prevalence of cognitive impairments and confidence interval (mild or MCI and severe or dementia)

|  |  | Prevalence<br>% | Lower<br>Bound | Upper<br>Bound |
| --- | --- | --- | --- | --- |
| HRS | Normal | 66.2 | 64.2 | 68.1 |
| US | MCI | 23.1 | 21.5 | 24.9 |
|  | Dementia | 10.7 | 9.5 | 12.0 |
| ELSA | Normal | 83.1 | 83.0 | 83.2 |
| England | MCI | 9.4 | 9.3 | 9.5 |
|  | Dementia | 7.5 | 7.4 | 7.5 |
| SHARE | Normal | 67.6 | 65.3 | 69.8 |
| Europe | MCI | 22.1 | 20.1 | 24.1 |
|  | Dementia | 10.4 | 9.0 | 11.9 |
| CHARLS | Normal | 93.7 | 93.0 | 94.2 |
| China | MCI | 6.2 | 5.6 | 6.8 |
|  | Dementia | 0.2 | 0.1 | 0.3 |

**Table 3.** Multinomial logistic regression of cognitive impairments (mild or MCI and severe or dementia) in US, China, England and Europe (pooled), N= 12862.

|  | MCI | Dementia |
| --- | --- | --- |
| Early life adversity | 0.215<sup>*</sup><br>(0.02) | 0.557<sup>***</sup><br>(0.00) |
| Age | 0.106<sup>***</sup><br>(0.00) | 0.0752<sup>***</sup><br>(0.00) |
| Sex, Female | -0.115<br>(0.09) | -0.0880<br>(0.49) |
| Poorest quartile | 0.507<sup>***</sup><br>(0.00) | 0.498<sup>***</sup><br>(0.00) |
| Some college | -0.986<sup>***</sup><br>(0.00) | -0.615<sup>***</sup><br>(0.00) |
| Urban | 0.843<sup>***</sup><br>(0.00) | 0.378<sup>**</sup><br>(0.00) |
| Married/union | -0.0656<br>(0.36) | -0.0801<br>(0.54) |
| Depression | 0.247<sup>**</sup><br>(0.00) | 0.871<sup>***</sup><br>(0.00) |
| US, ref. |  |  |
| England | -0.780<sup>***</sup><br>(0.00) | 0.289<br>(0.17) |
| Europe | -0.121<br>(0.25) | 0.110<br>(0.50) |
| China | -1.460<sup>***</sup><br>(0.00) | -3.647<sup>***</sup><br>(0.00) |
| Constant | -9.152<sup>***</sup><br>(0.00) | -8.080<sup>***</sup><br>(0.00) |

*p*-values in parentheses
<sup>\*</sup> *p* < 0.05, <sup>\*\*</sup> *p* < 0.01, <sup>\*\*\*</sup> *p* < 0.001

**Figure 2.**
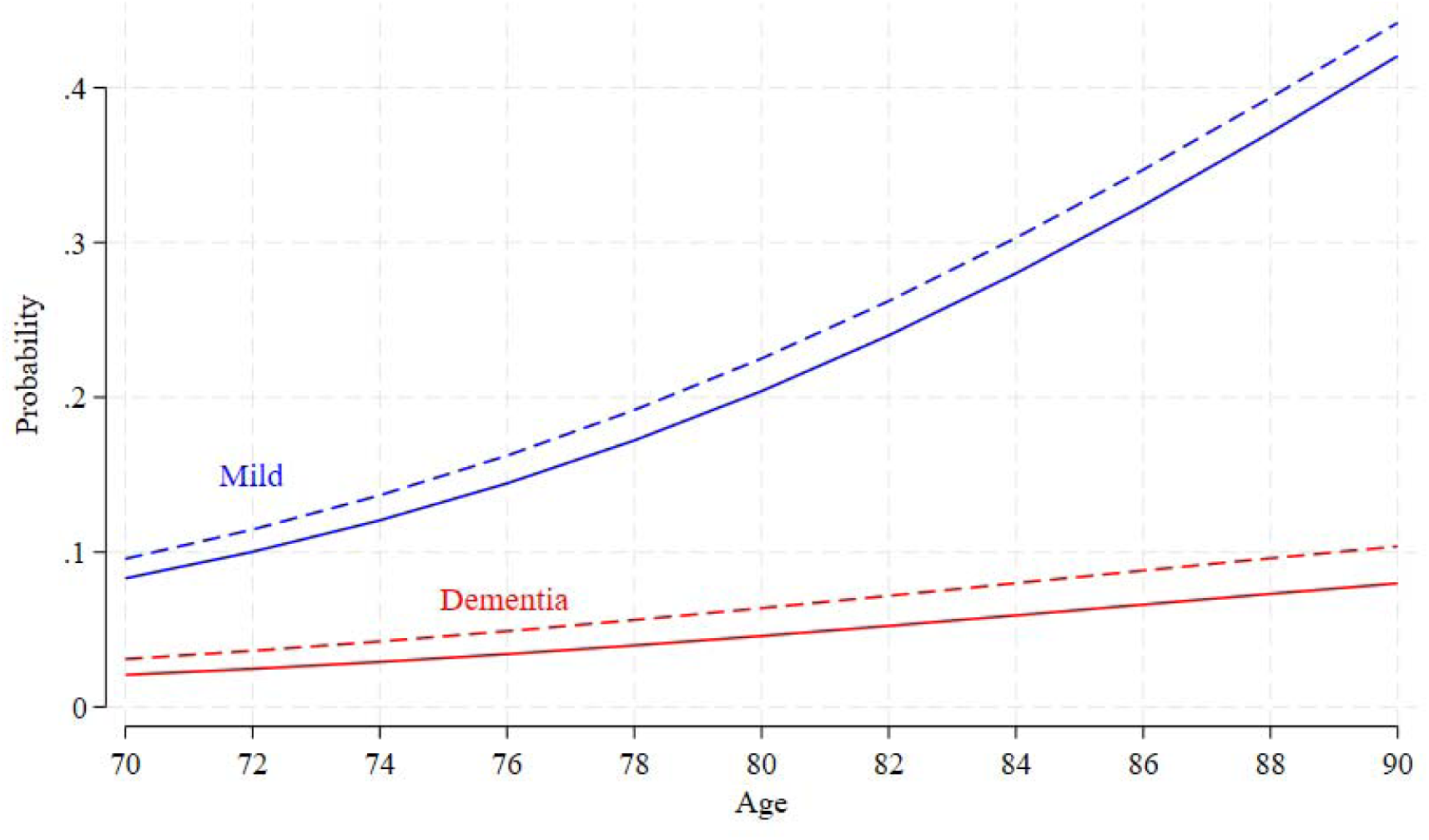
Marginal plot of probabilities of mild cognitive impairment (MCI in blue) and dementia (red) in adults aged 70 – 90 years, distinguishing the childhood poor (dash) from the non-poor (solid), pooled sample of US, China, England and Europe

We plotted marginal probabilities of MCI and dementia, distinguishing the childhood poor (dashed lines) from the non-poor (solid lines). Recall that cognitive status is nominal, which implies that dementia is directly compared to normal, likewise with MCI because not all MCI necessarily converts or progresses to dementia.^32^ See the supplement for an ordinal cognitive status and for combining MCI and dementia. The plot shows that age is associated with the probabilities of MCI and dementia in non-linear fashion especially of MCI (significance or p-values are presented in table 3). As age advances, so do the probabilities of MCI and dementia. On top of this, childhood poverty is significantly deleterious; dashed lines are always above solid lines.

We then rerun the multinomial regression for each survey (table 4) which shows that contrary to the results for the pooled sample, in the US and China childhood poverty does not significantly associate with MCI or dementia. But the results for England and Europe mirror the pooled results. In the event we then pooled England and Europe samples to re-estimate the associations and plot the marginal probabilities (table 5).

**Table 4.** Multinomial logistic regression of cognitive impairments (mild and dementia) separately in US (HRS), China (CHARLS), England (ELSA) and Europe (SHARE)

|  | HRS | N=1274 | CHARLS | N=8850 | ELSA | N=817 | SHARE | N=1921 |
| --- | --- | --- | --- | --- | --- | --- | --- | --- |
|  | Mild | Dementia | Mild | Dementia | Mild | Dementia | Mild | Dementia |
| Early life adversity | 0.134<br>(0.50) | 0.162<br>(0.63) | -0.138<br>(0.65) | 0.643<br>(0.48) | 0.956***<br>(0.00) | 0.567<br>(0.05) | 0.359*<br>(0.01) | 0.527**<br>(0.01) |
| Age | -0.239<br>(0.18) | -0.380<br>(0.17) | 6.503***<br>(0.00) | 26.48***<br>(0.00) | 0.474<br>(0.29) | 0.111<br>(0.79) | 0.00409<br>(0.19) | 0.00735*<br>(0.03) |
| Age-squared | 0.0017<br>(0.13) | 0.0029<br>(0.10) | -0.0361***<br>(0.00) | -0.144***<br>(0.00) | -0.0034<br>(0.32) | -0.0006<br>(0.86) | 0.0001<br>(0.30) | 0.0004***<br>(0.00) |
| Sex, Female | 0.0454<br>(0.77) | 0.0596<br>(0.83) | 1.132***<br>(0.00) | 3.329**<br>(0.00) | 0.130<br>(0.66) | -0.0510<br>(0.86) | -0.316**<br>(0.01) | -0.377*<br>(0.04) |
| Poorest quartile | 0.420<br>(0.08) | 0.646<br>(0.08) | 0.268<br>(0.15) | 1.657<br>(0.08) | 0.428<br>(0.16) | 0.717*<br>(0.01) | 0.744***<br>(0.00) | 0.557**<br>(0.00) |
| Some college | -0.0572<br>(0.69) | 0.0969<br>(0.70) | -4.899***<br>(0.00) | -19.59<br>(0.99) | -0.811<br>(0.07) | -0.0019<br>(1.00) | -0.0771<br>(0.58) | -0.724**<br>(0.01) |
| Urban | -0.204<br>(0.15) | -0.312<br>(0.21) | 7.699***<br>(0.00) | 27.69<br>(1.00) | 0.0399<br>(0.91) | -0.0033<br>(0.99) | -0.0690<br>(0.57) | 0.270<br>(0.15) |
| Married/union | -0.0147<br>(0.93) | -0.135<br>(0.64) | -0.137<br>(0.49) | 0.263<br>(0.79) | 0.134<br>(0.67) | -0.135<br>(0.65) | 0.366**<br>(0.00) | 0.456*<br>(0.02) |
| Depression | 0.654***<br>(0.00) | 0.467<br>(0.17) | 0.262<br>(0.26) | 0.294<br>(0.82) | 1.112***<br>(0.00) | 0.452<br>(0.17) | 0.435***<br>(0.00) | 1.372***<br>(0.00) |
| Constant | 6.769<br>(0.33) | 9.474<br>(0.38) | -294.6***<br>(0.00) | -1239.7<br>(0.82) | -19.41<br>(0.18) | -7.478<br>(0.58) | -2.092***<br>(0.00) | -5.660***<br>(0.00) |

*p*-values in parentheses \* *p* < 0.05, \*\* *p* < 0.01, \*\*\* *p* < 0.001

**Table 5.** Multinomial logistic regression of cognitive impairments (mild and dementia) separately in England (ELSA) and Europe (SHARE) (N = 2738)

|  | Mild | Dementia |
| --- | --- | --- |
| Early life adversity | 0.403**<br>(0.00) | 0.418**<br>(0.01) |
| Age | 0.005<br>(0.10) | 0.007*<br>(0.03) |
| Age-squared | 0.0001<br>(0.06) | 0.0004***<br>(0.00) |
| Sex, Female | -0.262*<br>(0.02) | -0.243<br>(0.10) |
| Poorest quart. | 0.433**<br>(0.00) | 0.427*<br>(0.01) |
| Some college | -0.160<br>(0.22) | -0.285<br>(0.14) |
| Urban | -0.091<br>(0.42) | 0.066<br>(0.67) |
| Married/union | 0.227*<br>(0.05) | 0.193<br>(0.20) |
| Depression | 0.481***<br>(0.00) | 1.033***<br>(0.00) |
| England, ref. |  |  |
| Germany | 0.637<sup>***</sup><br>(0.00) | -1.218<sup>***</sup><br>(0.00) |
| Italy | 0.661<sup>***</sup><br>(0.00) | -0.226<br>(0.29) |
| France | 0.345<br>(0.08) | -0.938<sup>***</sup><br>(0.00) |
| Denmark | 0.580<sup>**</sup><br>(0.00) | -0.553<sup>*</sup><br>(0.02) |
| Czechia | 1.050<sup>***</sup><br>(0.00) | -0.457<br>(0.06) |
| Constant | -2.885<sup>***</sup><br>(0.00) | -4.424<sup>***</sup><br>(0.00) |

*p*-values in parentheses
<sup>\*</sup> *p* < 0.05, <sup>\*\*</sup> *p* < 0.01, <sup>\*\*\*</sup> *p* < 0.001

**Figure 3.**
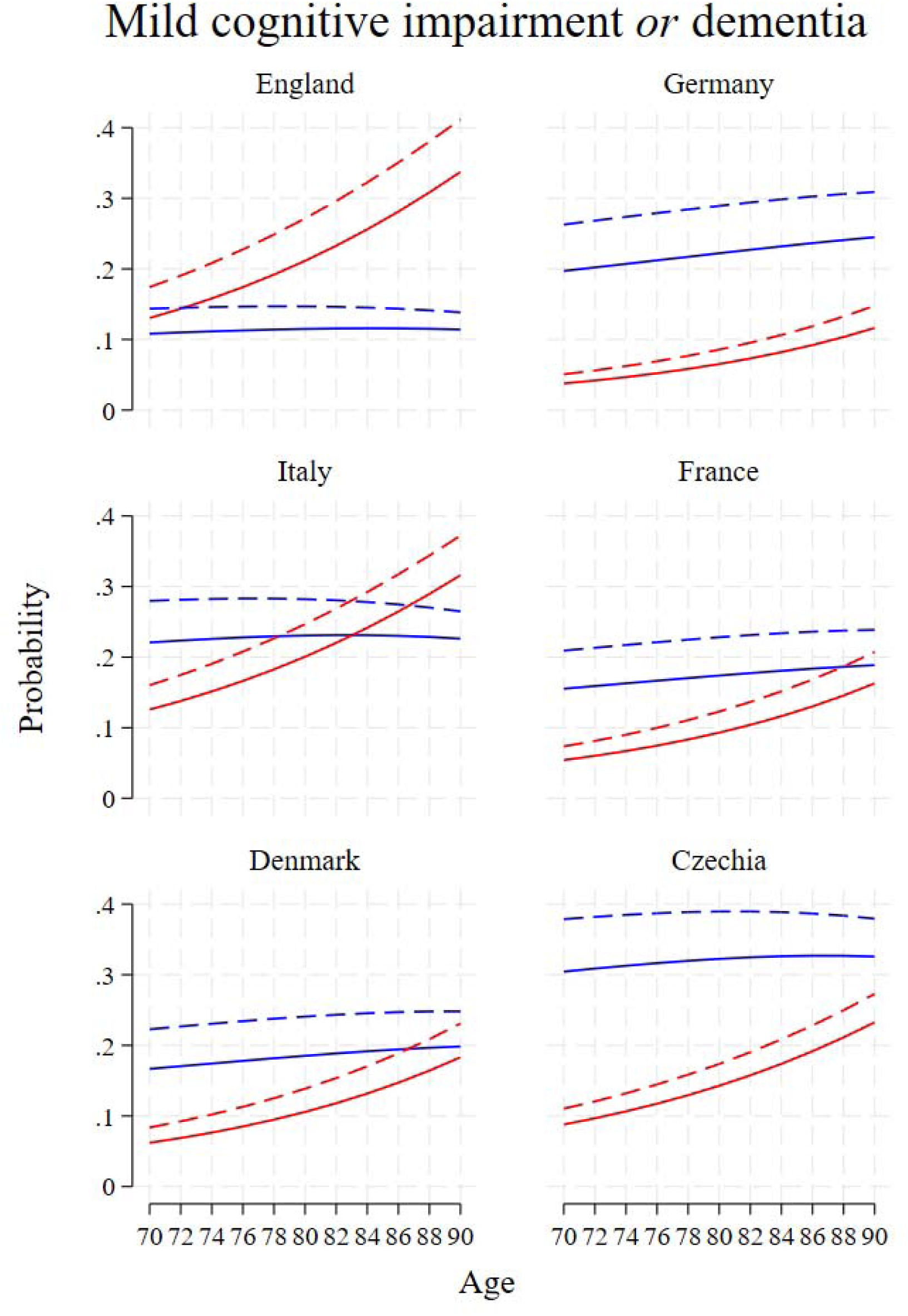
Marginal probabilities of MCI (blue) and dementia (red) in England and European countries, distinguishing the childhood poor (dash) from the non-poor (solid)

There are important similarities and differences worth noting between this set of estimates and plots and that of the pooled sample. Childhood adversity is significantly associated with MCI and dementia. The patterns of marginal probabilities in England across age and childhood adversity mimic the patterns for the pooled sample. This is remarkable because as we noted above the two largest surveys (US and China) are not used or did not show significant childhood adversity. The significance of childhood adversity as depicted in the distance of dashed lines above solid lines is evident in England and European countries. Some (though not exhaustive) comparative observations can be made. For example, on the right column and dementia probabilities – there is an increase in probabilities along this sequence of Germany, France and Czechia. Similarly on the left column along this sequence of Denmark, England and Italy. Altogether this is showing the well-known concentric pattern of European advantage: Scandinavian countries, Western European, Eastern or Southern European. For examples, our work showed this Scandinavian advantage with regards to frailty and to visual and hearing loss, two of the largest modifiable risk factors identified in the commission’s life course frame above.^9,38^

Briefly, the sensitivity analyses where cognitive status was entered as ordinal and as binary show the main results to be robust: early life adversity is associated with late life cognitive impairment; see the supplement.

## DISCUSSION

The life course hypothesis posits that early life adversity continues to be associated with cognitive impairment (MCI and dementia) in late life irrespective of risk factors in adulthood such as education. Thanks to ongoing efforts at harmonisation, for the first time we found evidence largely in its support across wealthy nations, echoing recent global evidence on the life course shaping of frailty and multimorbidity.^8–10^

We further discuss key results spanning substantive and methodological aspects. First, the prevalence rates of dementia vary appreciably, from 0.2% (China) to 10.7% (US). Among the rich countries the rates are evidently similar. This variation is not attributable to measurement or protocol. The observed variation highlights the influence of social, historical, and policy contexts across countries, suggesting that national differences in healthcare systems, social support, and policy interventions may contribute to cognitive outcomes.^39,40^ An additional driver behind the variation is the self-selection of those responding to the harmonisation project which may have led to different prevalence rates.^2,4,23,41^ Nevertheless, this new study demonstrates the advantage of harmonisation especially in estimating the strength of association which is our aim. A similar focus on association (not on prevalence) has been called for when investigating depression and allostatic load in ageing populations.^39,42^

Second, early life adversity is associated with mild cognitive impairment and with dementia in complex ways. In Europe and England, the childhood poor are significantly at higher risk of cognitive impairment (MCI and dementia). The magnitudes are also considerable. Not so in America and China where early life adversity is not significantly associated with either MCI or dementia. It is not possible to give a definitive answer as to why this is so. Nonetheless overlapping explanations can be suggested.

In the case of China, selective mortality or, conversely, survival may play a role. Half a century ago China was largely poor and such poverty may have contributed to more deaths among the poor, leaving as survivors those who are uniquely advantaged including genetically. In this way the sample has been selected by decades of selective survival.^14,24^ Thus early life adversity no longer matters. This may also lie behind the null result in the U.S.

And a cross-country design offers an enrichment to this explanation. Until recently healthcare has been largely private in America, often tied to workplaces, exposing millions without jobs to the vagaries of chance.^43,44^ Effectively, the same process of selection is operating there over a long period of life until they are eligible for Medicare. In comparison with nations across the Atlantic which tend to have public or social insurance health care, e.g. the English National Health Service, the HRS sample was exposed to non-negligible selective mortality.^14,43^ Health policy is not the only driver separating both sides of the Atlantic; history is also a factor. On depression and cognitive trajectories, for example, we showed that World War II was decisive in changing the cohort composition of older populations.^40,45^ Taken together, these may drive the US results.

Having discussed substantive findings, we reflect on methodology (retaining the numbering). The third illustrates the importance of a latent construct for cross-country harmonisation of cognitive status. The HCAP network deployed multiple instruments with differences adjusted to the national contexts, from the obvious ones (Prime Minister instead of President) to the more elaborate cultural stories. Also, the number of items varies.^3,19,20,24^ The success of this harmonisation critically depends on the use of latent constructs, specifically confirmatory (latent) factor analysis. Without the latent construct there is no threefold cognitive status.

In parallel, without the latent construct there is no binary early life adversity for testing the life course hypothesis. We have used latent construction, specifically confirmatory latent class analysis, to obtain childhood adversity status and explain sarcopenia, depression, episodic memory, frailty and multimorbidity in up to 31 countries both wealthy and developing.^7–10^ The symmetry of harmonisation of cognitive status and of adversity status is made transparent, a methodological step that relies critically on latent constructs. Thereby, the tests of the life course hypothesis around the world are made possible and strong. Otherwise it would be difficult to make headway in rectifying the omission of early life adversity from the commission’s life course frame on global samples. By symmetrically treating late life outcome and early life adversity, the life-course shaping hypothesis can be reliably tested.

This fruit of parallel use of latent constructs has further significance. This family of studies is growing to other countries in different countries or regions, e.g. Ireland, South America or Southeast Asia, as broadcast through the <u>HCAP network</u> (accessed 28 May 2025). With different indicators of adversity and different indicators of outcome in different world regions, a latent construction of both early life adversity and late life outcome is a viable methodology to study the life course shaping of health with retrospective information. This, in turn, directly helps in bringing along every older adult in achieving a healthy life as is the aspiration of the UN Decade of Healthy Ageing. Internationally comparable cognitive status and its association with life course risk factors are critical to this global initiative, for without them global monitoring will be curtailed.

This study has strengths and weaknesses and noting them is important for the ongoing longitudinal ageing studies and their harmonisation. A key strength lies in the breadth of coverage which, according to the <u>UN World Population Prospects 2024</u>, encompasses regions representing 31.9% of the global population and 50.4% of the older population (60 and over) (accessed 1 June 2025). This expansive coverage allows for meaningful cross-country comparisons between diverse socioeconomic and healthcare systems, including both wealthy and developing nations. Second, the key exposure and outcome are harmonised which made interpretation for all countries in the sample immediate. Last, the same method is modular, easily deployed when for instance Ireland or the Irish Longitudinal Study on Ageing released its HCAP data.

The weaknesses are several. First, this is not a randomised experiment study, thus precluding causal inference. Note that we discussed various issues of randomised experiment for such a long time separation between the posited cause (early life adversity) and an outcome (older age cognition).^8^ Second, the association between childhood and late life cognition is direct and not mediated. A study on SHARE for instance used the same sample to unpack the association between childhood adversity and late life self-rated health by including adolescent health midway between the risk factor and the outcome in a structural equation model.^17^ Note however that this type of structural equation model is not applicable when the childhood adversity indicators vary across surveys (recall the discussion on starvation during China’s Great Leap Forward above). This is left for future work. Third, cognitive status in these countries was not ascertained by clinicians. Nevertheless, the commission also based their evidence on similar materials, including our own work.^31^ Fourth, it has not done full justice to the commission’s life course frame above, due to lack of risk factors such as traumatic brain injury and untreated visual loss in all of the ageing surveys.^29,30^

Last, cross-country research design necessarily sacrifices what is unique about each country, not least its social norm about the role of older adults embedded in communities which may shape social isolation, an important modifiable risk factor according to the commission. Treatment of each country in this group must wait for a monograph to do it justice, along the lines of Simone de Beauvoir’s major work on ageing (1972/1996).

In conclusion, early life adversity is associated both with mild cognitive impairment and dementia in many wealthy nations. In a middle-income nation like China, other risk factors along the life course may have exerted their influence earlier and more effectively. This complexity in the shaping of health in late life is revealed for the first time thanks to the latent constructions of harmonised key risk factor and outcome. Future monitoring of global initiatives such as the UN Decade of Healthy Ageing can employ the methodology more widely, specifically to the expanding network of harmonised cognitive assessment protocol such as the Irish Longitudinal Study on Ageing. Future attribution of dementia and the application of the life course frame can benefit from recognising the lifelong reach of early life adversity.

## ETHICAL REVIEW

The University of Manchester exempted the investigation from full ethical review as it uses publicly available deidentified secondary datasets.

## AUTHOR CONTRIBUTIONS

The corresponding author has read the policies on author responsibilities and submits this manuscript in accordance with those policies.

## ACKNOWLEDGEMENT

We thank the participants in eight countries for providing information, time and in many visits blood biomarkers too, as well as the generous funding bodies over many decades. **HRS**: The HRS (Health and Retirement Study) is sponsored by the National Institute on Aging (grant number NIA U01AG009740) and is conducted by the University of Michigan. **ELSA**: The English Longitudinal Study of Ageing was developed by a team of researchers based at University College London, NatCen Social Research, the Institute for Fiscal Studies, the University of Manchester and the University of East Anglia. The data were collected by NatCen Social Research. The funding is currently provided by the National Institute on Aging in the US (2RO1AG7644 and 2RO1AG017644-01A1), and a consortium of UK government departments coordinated by the National Institute for Health Research. **SHARE**: This paper uses data from SHARE Waves 3, 7 and 9 (DOIs: 10.6103/SHARE.w3.800, 10.6103/SHARE.w7.800, 10.6103/SHARE.w9ca800) see Börsch-Supan et al. (2013) for methodological details. The SHARE data collection has been funded by the European Commission, DG RTD through FP5 (QLK6-CT-2001-00360), FP6 (SHARE-I3: RII-CT-2006-062193, COMPARE: CIT5-CT-2005-028857, SHARELIFE: CIT4-CT-2006-028812), FP7 (SHARE-PREP: GA N°211909, SHARE-LEAP: GA N°227822, SHARE M4: GA N°261982, DASISH: GA N°283646) and Horizon 2020 (SHARE-DEV3: GA N°676536, SHARE-COHESION: GA N°870628, SERISS: GA N°654221, SSHOC: GA N°823782, SHARE-COVID19: GA N°101015924) and by DG Employment, Social Affairs & Inclusion through VS 2015/0195, VS 2016/0135, VS 2018/0285, VS 2019/0332, and VS 2020/0313. Additional funding from the German Ministry of Education and Research, the Max Planck Society for the Advancement of Science, the U.S. National Institute on Aging (U01_AG09740-13S2, P01_AG005842, P01_AG08291, P30_AG12815, R21_AG025169, Y1-AG-4553-01, IAG_BSR06-11, OGHA_04-064, HHSN271201300071C, RAG052527A) and from various national funding sources is gratefully acknowledged (see www.share-project.org).

## DATA AVAILABILITY

HRS, CHARLS, SHARE and ELSA are freely available to researchers. Access can be obtained after registration with respective repositories (HRS) https://hrs.isr.umich.edu, (SHARE) www.share-project.org, (CHARLS) https://charls.pku.edu.cn, and (ELSA) www.elsa-project.ac.uk.

## SUPPLEMENT

Early life adversity and late life cognitive impairment (any impairment: MCI plus dementia)

**Figure supplement 1.**
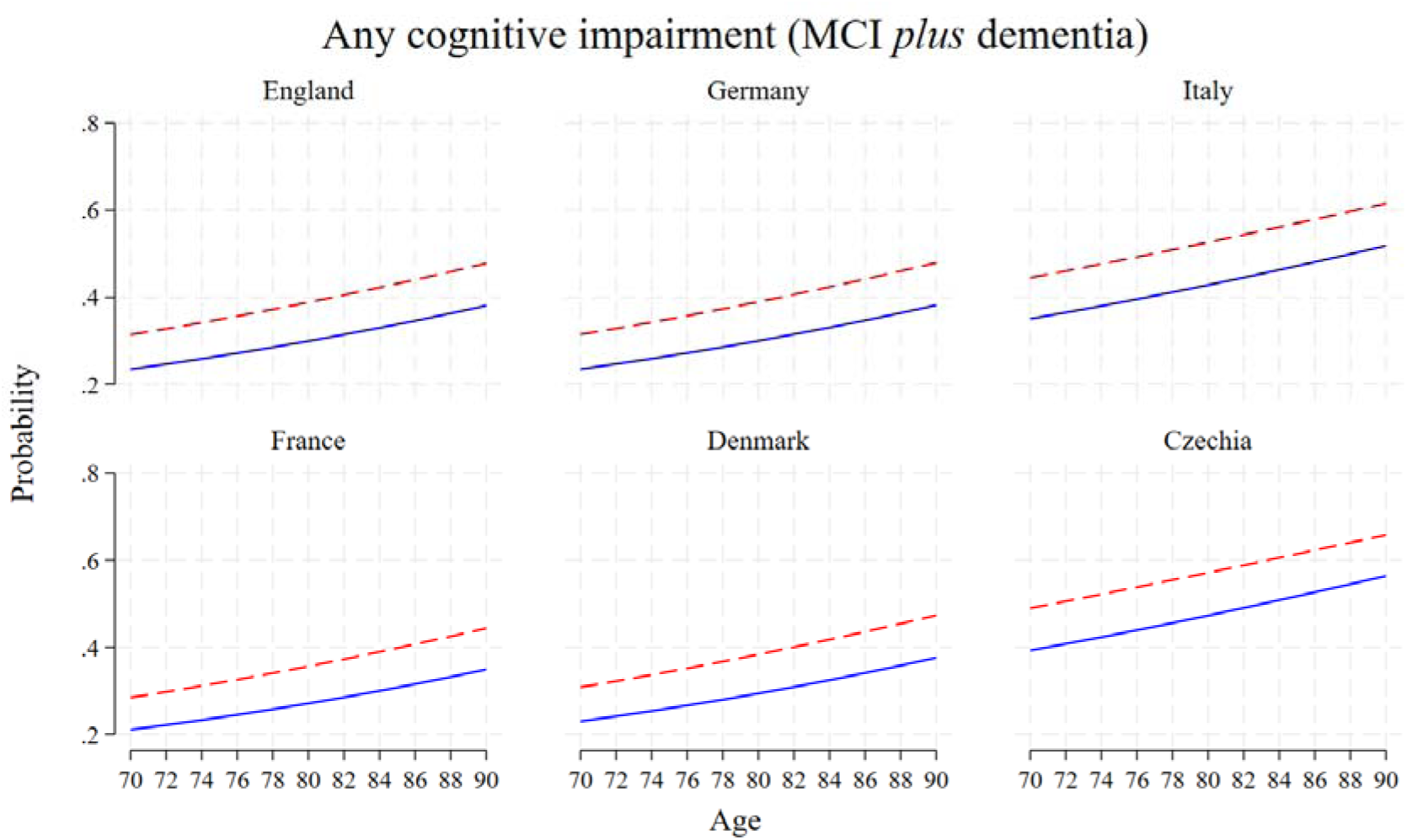

**Table supplement 1.** Logistic regression any cognitive impairment (MCI plus dementia)

|  | Any cognitive<br>impairment |
| --- | --- |
| Early life adversity | 0.410***<br>(0.00) |
| Age | 0.006*<br>(0.02) |
| Age-squared | 0.0002***<br>(0.00) |
| Sex, Female | -0.258**<br>(0.01) |
| Poorest quart. | 0.432***<br>(0.00) |
| Some college | -0.200<br>(0.08) |
| Urban | -0.0398<br>(0.68) |
| Married/union | 0.211*<br>(0.03) |
| Depression | 0.676***<br>(0.00) |
| England, ref. |  |
| Germany | 0.0481<br>(0.75) |
| Italy | 0.320*<br>(0.04) |
| France | -0.126<br>(0.44) |
| Denmark | 0.153<br>(0.33) |
| Czechia | 0.517***<br>(0.00) |
| Constant | -2.671***<br>(0.00) |
| *N* | 2738 |

*p*-values in parentheses
\* $p < 0.05$ , \*\* $p < 0.01$ , \*\*\* $p < 0.001$

**Figure Supplement 2.**
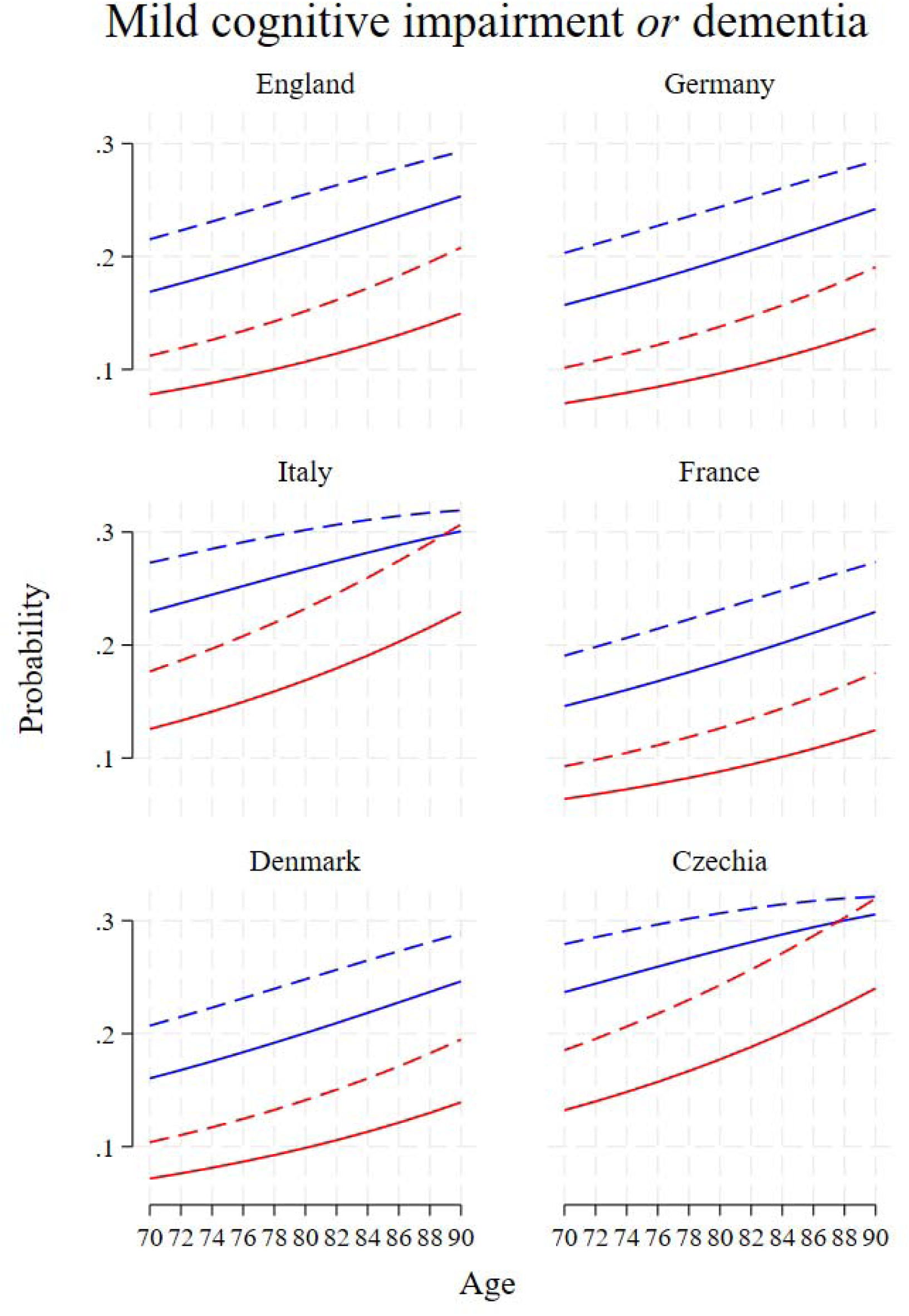

**Table Supplement 2.**

|  |  |
| --- | --- |
| Early life<br>adversity | 0.410<sup>***</sup><br>(0.00) |
| Age | 0.005<sup>*</sup><br>(0.03) |
| Age-squared | 0.0002<sup>***</sup><br>(0.00) |
| Sex, Female | -0.241<sup>**</sup><br>(0.01) |
| Poorest quart. | 0.414<sup>***</sup><br>(0.00) |
| Some college | -0.196<br>(0.08) |
| Urban | -0.0198<br>(0.84) |
| Married/union | 0.188<br>(0.05) |
| Depression | 0.724<sup>***</sup><br>(0.00) |
| England, ref. |  |
| Germany | -0.0674<br>(0.65) |
| Italy | 0.269<br>(0.07) |
| France | -0.206<br>(0.20) |
| Denmark | 0.0945<br>(0.54) |
| Czechia | 0.346<sup>*</sup><br>(0.02) |
| Threshold 1 | 2.657<sup>***</sup><br>(0.00) |
| Threshold 2 | 4.042<sup>***</sup><br>(0.00) |
| *N* | 2738 |

*p*-values in parentheses
<sup>\*</sup> *p* < 0.05, <sup>\*\*</sup> *p* < 0.01, <sup>\*\*\*</sup> *p* < 0.001

